# Data-driven robust machine learning models to differentiate Parkinson’s disease patients using heterogeneous risk factors

**DOI:** 10.64898/2025.12.18.25342612

**Authors:** Malinda Iluppangama, Dilmi Abeywardana, Chris P. Tsokos

**Affiliations:** Department of Mathematics and Statistics, Loyola University Maryland, Baltimore, MD, USA; Department of Mathematics and Statistics, University of South Florida, Tampa, FL, USA

## Abstract

Parkinson’s Disease (PD) is the most prevalent neurodegenerative disorder after Alzheimer’s, yet its diagnosis is largely based on subjective clinical assessments. Thus, this comparative study proposes a systematic, data-driven approach to accurately classify PD patients using heterogeneous risk factors along with efficient machine learning models. Six machine learning algorithms, Support Vector Machine(SVM), Random Forest(RF), Extreme Gradient Boosting(XGBoost), Logistic Regression(LR), K-Nearest Neighbor (KNN), and Decision Tree(DT), were utilized and evaluated to identify the robust and efficient model with high discrimination power. The SVM model outperformed all other machine learning models, and it has been identified as a robust model to classify PD patients from healthy individuals with a 98% accuracy based on training phase performance. Furthermore, feature importance was analyzed using SHAP to enhance the interpretability of the proposed model. This study contributes to the growing use of artificial intelligence in healthcare by exploring data-driven classification methods. These models may help support healthcare professionals by providing additional information for identifying high-risk patients. Our results suggest that these approaches may help improve early detection of PD, with proper validation.

## 1 Introduction

The nervous system, including the brain, spinal cord, and other nerves, plays a critical role in controlling movement, cognition, and behavior. Damage, dysfunction, or abnormalities in nerves system can lead to a range of symptoms that affecting movement, thinking, and sensory processes. These conditions classified as neurological disorders. Neurological disorders represent a global health burden and a leading cause of disability, and the second largest cause of death in the world. In 2019 they were responsible for nearly 10 million deaths globally, [1], [2]. These disorders can be broadly classified into neurotraumatic diseases(i.e., spinal cord injury, stroke, and traumatic brain injury), neurodegenerative diseases (Alzheimer’s disease and Parkinson’s disease (PD)), and neuropsychiatric diseases (major depression, tardive dyskinesia, and epilepsy), [3].

This study focuses primarily on neurodegenerative disorders. Gradual deterioration and death, as well as structural changes and the inability of neurons inside the brain and central nervous system to function normally, are known as neurodegeneration. Alzheimer’s disease is the largest neurodegenerative disease worldwide and eventually leads to the **progressive neurodegenerative condition of dementia**, while PD ranks as the second largest neurodegenerative disorder, [4], [5]. In this study, our main focus is on PD, which is associated with significant motor and non-motor symptoms, including potential cognitive decline.

PD is the second largest chronic neurodegenerative condition affecting the neuronal system of the brain after Alzheimer’s disease worldwide. Dopaminergic neurons are a type of neuron located in the substantia nigra, and they produce and release the neurotransmitter dopamine. These neurons are the main source of dopamine in the midbrain, and dopamine controls body movements as well as behavioral process such as mood, stress, and addiction. A certain level of dopamine is essential for normal human movements. If these dopaminergic neurons either overproduce or underproduce dopamine, it can lead to movement disorders, [6]. If these neurons suddenly stop functioning, die, or undergo structural changes, it can result in a shortage of dopamine in the midbrain. As a result, the body loses control of its movements, and medical professionals diagnose this condition as PD, [7], [8]. Fig 1 compares the healthy neuron with a neuron affected by PD, showing that healthy neurons have a higher dopamine concentration than the PD-affected neuron.

**Fig 1.**
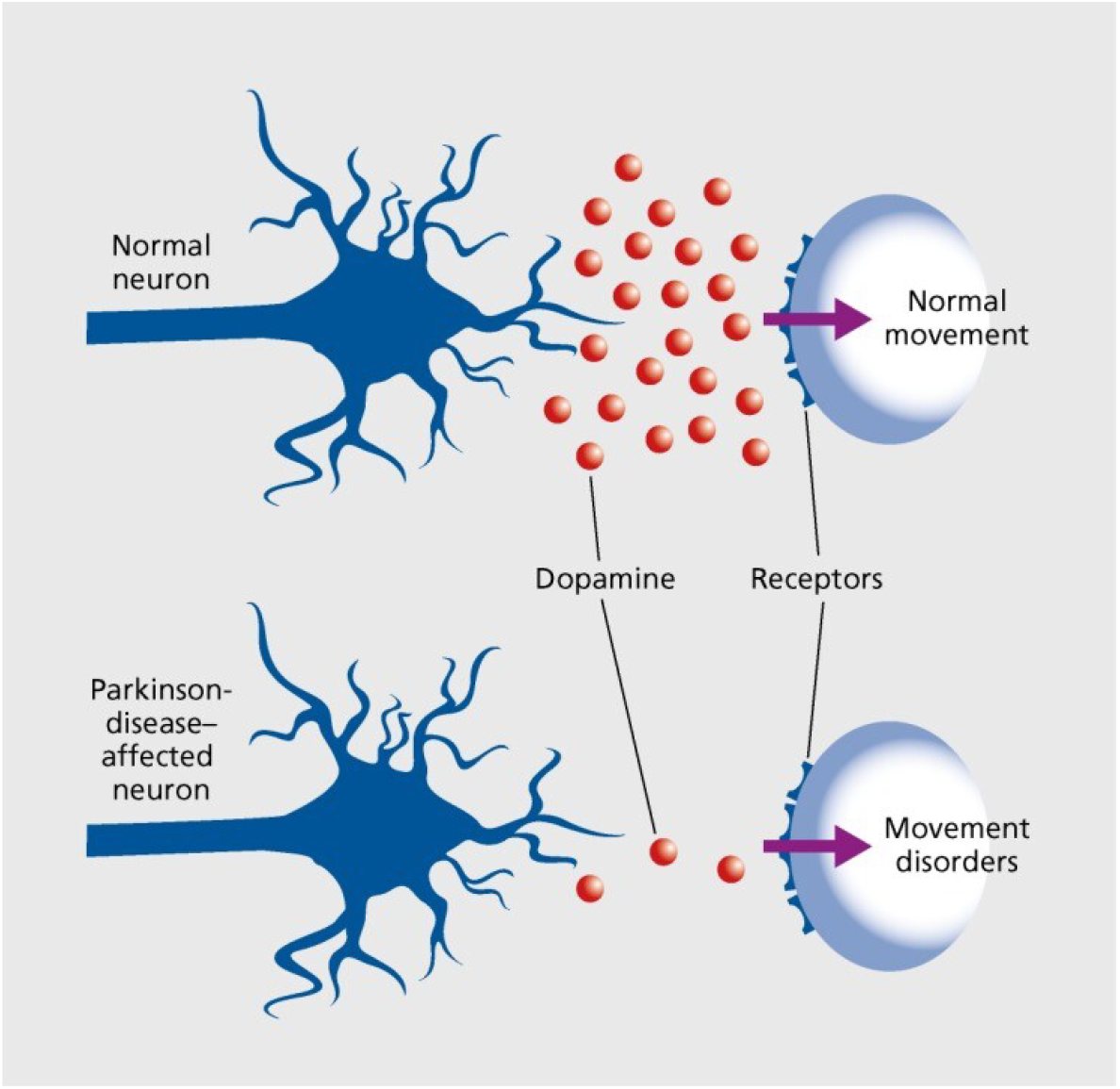
Comparison of Healthy Neuron Vs. Neuron Affected with PD

There are more than 10 million people worldwide living with this **severe neurodegenerative disorder**. For example, about one million people are diagnosed with PD in the USA alone, and approximately 90,000 new cases are identified each year. These numbers do not reflect the thousands of cases that remain undetected because some people live with PD, being aware of it. Furthermore, the number of PD patients is expected to reach 1.2 million by 2030. These figures highlight the serious global impact of this **neurodegenerative disorder** and the urgent need to address it to improve the quality of life to those living with PD, [9], [10].

According to medical professionals, PD can be classified into three major categories: young-onset PD, genetic PD, and idiopathic PD. Idiopathic PD is the most common type, while the other two forms account for approximately 10% to 15% of total PD cases worldwide. Therefore, this study, focuses on idiopathic PD. Tremor, Bradykinesia, Stiffness, and Walking and Balance problems are the four cardinal symptoms used by medical professionals to diagnose idiopathic PD. These symptoms worsen over time, and vary among patient. Therefore, it is difficult for medical professionals to predict the symptoms that a particular patient may develop in the future, [11], [12], [13]. Furthermore, no cure for PD has been identified to date. However, several medications, such as Levodopa, Carbidopa, and dopamine agonists, along with therapies such as focused ultrasound and surgical procedures like deep brain stimulation, have been introduced to slow disease progression and improve quality of life, [14]. **Scientists have increasingly focused on understanding the mechanisms behind PD and developing effective treatments to reduce its burden on patients, families, and healthcare systems**, [15], [16], [17], [18], [19]. In addition to individual researchers, there are several research centers and foundations such as the Parkinson’s Foundation, the Michael J. Fox Foundation for Parkinson’s Disease and the National Institute of Health, contribute to the study of PD and support efforts to better understand the disease and develop potential treatments.

Although extensive research has been conducted on PD and related disorders, formal diagnostic tests such as imaging or blood tests are not yet available. Moreover, the etiology of PD remains unclear, and medical professionals rely on descriptive methods to distinguish PD patients from healthy individuals. further research is essential to uncover valuable insights into this,critical disease and to support the development of new therapies and treatments, ultimately improving the lives of patients with PD.

Artificial intelligence (AI) represents the state of the art of many data-driven models across domains such as engineering, health sciences, cybersecurity, finance, environmental science, and has been successfully applied in a wide range of applications within these areas, [20], [21], [22]. AI is often associated with the concept of big data. However, in the domain of health science, obtained large datasets is challenging due to privacy concerns and the high cost of data collection, [23], [24]. Despite these limitations, researchers have effectively applied AI approaches particularly machine learning and deep learning, even with limited data, [25], [26], [27]. Among these approaches, supervised learning is a powerful subset of AI that is widely used by practitioners to develop various models to addressing problems in health science, [28], [29].

In recent years, machine learning and deep learning techniques have been increasingly employed to improve the understanding and diagnosis of Parkinson’s disease (PD), [30]. Among these approaches, Support Vector Machines (SVMs) are commonly used due to their strong classification capabilities. For example, one study combined SVM with high-dimensional radiomic features to differentiate PD patients from healthy controls, achieving an accuracy of approximately 90% across two independent test datasets, [31]. Another study integrated multimodal MRI data with cerebrospinal fluid biomarkers and developed an automated SVM-based model for early PD detection, reporting an accuracy of around 86%, [32]. In addition to MRI-based methods, researchers have also explored features derived from FP-CIT SPECT imaging using data from the Parkinson’s Progression Markers Initiative (PPMI). In this context, classification models based on SVM, k-nearest neighbors (KNN), and logistic regression (LR) have shown strong performance, with features such as putaminal binding potential achieving classification accuracy close to 95%, [33]. More recently, advanced ensemble and deep learning approaches have been introduced. One study utilized PPMI data along with a combination of Extreme Gradient Boosting (XGBoost) and multilayer perceptron (MLP) models to distinguish among different cognitive stages of PD, including PD with normal cognition (PD-NC), PD with mild cognitive impairment (PD-MCI), and PD dementia (PDD). Furthermore, SHapley Additive exPlanations (SHAP) were applied to improve model interpretability, and the proposed approach achieved a multi-class classification performance with an AUC exceeding 0.85, [34].

Despite the results of these prior studies, several limitations remain. Many previous studies focus on a limited number of machine learning models or depend on a single data modality, such as imaging or biomarkers, which might reduce the generalizability and robustness of the models. Additionally, a few studies reported comparative analysis of multiple machine learning models using a unified data set and consistent evaluation methods. To address these gaps in this study, we will develop a systematic comparative study to compare six well-defined machine learning models by utilizing multi-modal features, including features extracted from brain imaging, cerebrospinal fluid biomarkers, and potential genetic factors derived from the Parkinson’s Progression Markers Initiative (PPMI) dataset. Finally, the performance of these models is evaluated using rigorous cross-validation strategies and multiple evaluation metrics. This work aims to identify a reliable and robust approach for accurately distinguishing PD patients from healthy individuals.

The remainder of the paper is structured as follows. Section 2 presents the real dataset and describes the data pre-processing, including exploratory analysis using graphical methods and well-defined descriptive statistics. In Section 3, we develop six well-defined machine learning classification models and compare their performance to identify the most robust model for distinguishing patients with Parkinson’s disease (PD) from healthy individuals. This comparison is based on widely used evaluation metrics, including accuracy, area under the curve (AUC), precision, F-1 score, and recall. Finally, Section 4 concludes the chapter by summarizing the key findings, highlighting the most effective machine learning approach for discriminating PD patients from healthy individuals, and outlining directions for future research.

## 2 The Real Data

In developing and evaluating the proposed machine learning models, the present study obtained real data from the Parkinson’s Progression Markers Initiative (PPMI) through the Michael J. Fox Foundation. The dataset consists of information about 618 individuals, of whom 482 were diagnosed with PD, and the remaining 136 did not have PD.

From the literature review, we identified that clinical imaging features, including the left putamen, right putamen, left caudate, and right caudate, play a crucial role in distinguishing patients with Parkinson’s disease (PD) from healthy individuals. These brain region measurements are closely associated with dopaminergic activity and have been widely recognized as reliable biomarkers for PD detection in previous studies, [30], [35], [36], [37]. Furthermore, evidence suggests that a family history of PD significantly increases the risk of developing the disease, [38], [39]. In addition, biomarkers such as Tau, phosphorylated Tau (p-tau), Amyloid-beta, and *α*-Synuclein proteins provide valuable insights into the underlying mechanisms of neurodegenerative diseases, including PD. Abnormal concentrations of these proteins have been clinically linked to neuronal damage and disease progression, making them important candidates for early-stage PD classification, [40], [41]. After identifying these candidate variables from the peer-reviewed literature, we conducted a correlation analysis in combination with a purposeful variable selection approach to remove redundant and less informative features. As a result, seven key risk factors were selected as the most relevant and non-redundant predictors for developing data-driven machine learning models aimed at early PD detection, [42], [43], [44]. The dataset used in this study includes individuals aged from 30 to 85 years. It comprises several continuous risk factors, including concentrations of *β*-amyloid protein, p-tau protein, *α*-Synuclein, left caudate (CAUDATE*−*L), left putamen (PUTAMEN*−*L), and right putamen (PUTAMEN*−*R), and a categorical variable representing the individual’s family history. Fig 2, given below, illustrates the visualization of the data distribution,

**Fig 2.**
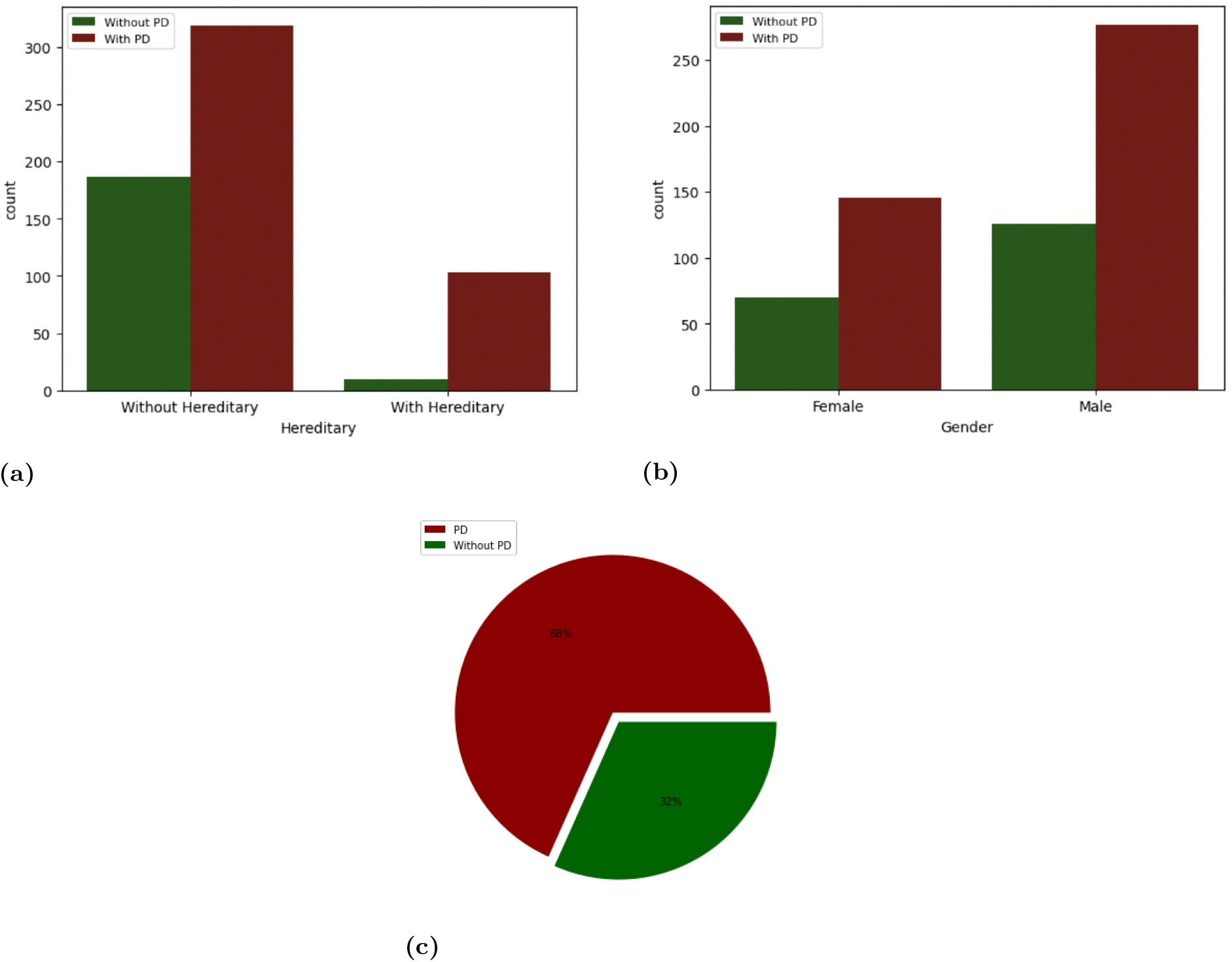
Data Visualization Across Possible Categories.

Table 1 presents the descriptive statistics of the key variables used to develop the machine learning models. To identify the relationships between each pair of individual variables, a correlation analysis was performed on the data. The heatmap of the correlation coefficients shown in Fig 3 illustrates the relationships among all possible pairs of explanatory variables.

**Table 1.** Descriptive Statistics of Continuous Risk Factors.

| Risk Factor | Minimum | Maximum | Mean | Standard Deviations |
| --- | --- | --- | --- | --- |
| β-protein | 238.80 | 3707.00 | 943.86 | 435.81 |
| p-tau protein | 8.01 | 73.61 | 15.71 | 6.16 |
| α-Synuclein | 432.40 | 5256.90 | 1565.22 | 689.69 |
| CAUDATE-L | 0.35 | 5.30 | 2.32 | 0.77 |
| PUTAMEN-R | 0.14 | 3.76 | 1.26 | 0.75 |
| PUTAMEN-L | 0.12 | 4.27 | 1.23 | 0.75 |

**Fig 3.**
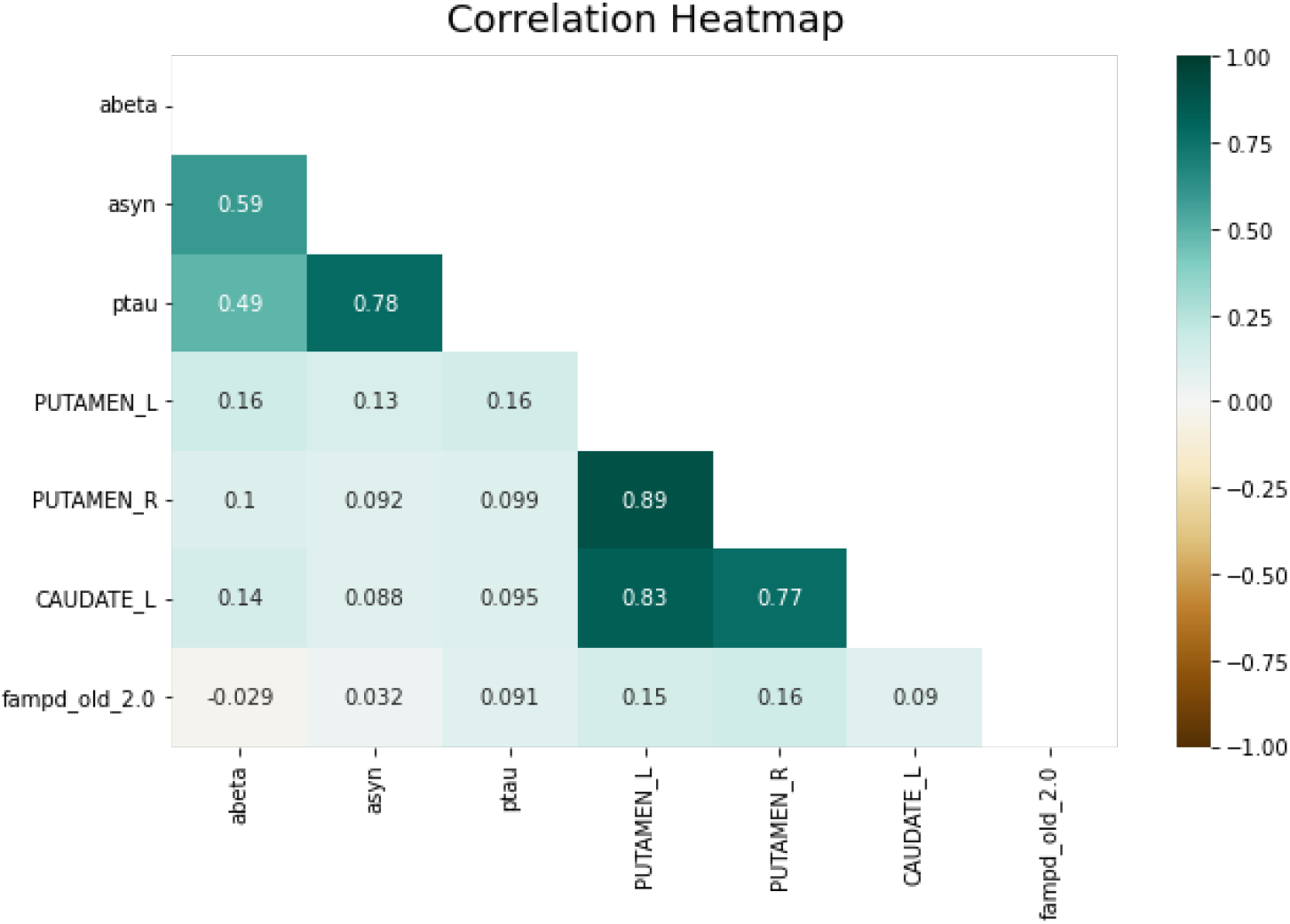
Correlation Coefficients of Risk Factors.

## 3 Model Development and Evaluation

In this study, the main objective is to accurately and efficiently distinguish between Parkinson’s disease (PD) patients and healthy individuals using six well-established artificial intelligence–based models, in order to identify a robust classification approach. Specifically, we employ six widely recognized supervised machine learning algorithms: Random Forest (RF), Extreme Gradient Boosting (XGBoost), Support Vector Machine (SVM), Decision Tree (DT), k-Nearest Neighbors (KNN), and Logistic Regression (LR). A brief overview of these methods is provided in the supplementary material,see S1 File, where the key concepts underlying model are summarized. The features selected from the PPMI dataset, described in Section 2, were used to develop these machine learning models to classify patients based on a given set of risk factors. Each model was evaluated using comprehensive performance metrics, including Area Under the Curve (AUC), accuracy, F-score, precision, and recall. A brief overview of these metrics is provided in the following section [45], [46], [47].

### 3.1 Performance Evaluation Framework

Model performance was evaluated using standard metrics derived from the confusion matrix. A confusion matrix summarizes classification results in terms of true positives (TP), true negatives (TN), false negatives (FN), and false positives (FP). A true positive and true negative occur when the predicted class matches the actual class for positive and negative cases, respectively, whereas false negatives and false positives represent misclassifications of positive and negative cases. Confusion matrix is summarized in Table 2,

**Table 2.** Confsusion Matrix.

|  |  | Predicted |  |
| --- | --- | --- | --- |
|  |  | True | False |
|  | True | **TP** | **FN** |
|  | False | **FP** | **TN** |

Table notes: TP=True Positive, TN=True Negative, FP= False Positive,FN=False Negative.

The confusion matrix based metrics were used to evaluate the performance of the proposed predictive models. Overall Accuracy, Precision, Recall, and F1-score were calculated from the confusion matrix to evaluate the performance of the propose models [48], [49]. A brief introduction on each of the evaluation metrics given below,

#### 3.1.1 Accuracy

The accuracy of a given model is the ability to identify true positive cases as true positive cases and true negative cases as true negative cases, which can be calculated by Eq 1,

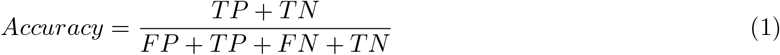

#### 3.1.2 Precision

Precision measures the proportion of true positives among all predicted positive cases and is calculated as given in Eq 2

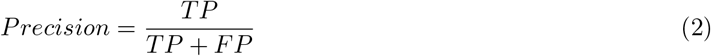

#### 3.1.3 Recall

Recall assesses a model’s ability to correctly identify positive instances. It is calculated using Eq. 3 and represents the proportion of true positives among all actual positives.

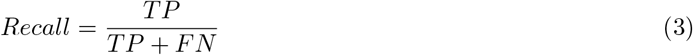

#### 3.1.4 F1-Score

The F1-score evaluates model performance by combining both precision and recall. It is calculated as the harmonic mean of these two measures, providing a balanced assessment of the model. The mathematical formulation of the F1-score is given in Eq. 4.

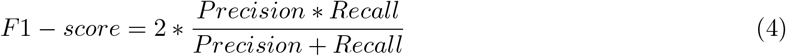

In addition to the confusion matrix, we evaluated the performance of the proposed models using Receiver Operating Characteristic (ROC) curves. ROC curves are created by plotting sensitivity against specificity for different classification thresholds. The area under the ROC curve (AUC) is used as a summary measure of model performance. A higher AUC value indicates better classification ability of the model [50].

### 3.2 Model Training and Evaluation

The model development procedure began with data preprocessing. First, the entire data set was split into two groups: train/validation and test. The train/validation set consists of 80% of the available data, while the remaining (20%) belongs to the test set. This split was performed before model training to ensure that the test dataset remains completely unseen during the model development process. Before model training, we evaluated the performances of all models using same evaluation scheme. However, the performance of some models was not satisfactory. Therefore, we decided to identify optimal hyperparameters to improve model performance through hyperparameter tuning using Grid Search Cross-Validation (GridSearchCV). This approach performs an extensive search over a predefined set of hyperparameter values, evaluates each combination using cross-validation, and selects the combination that maximizes validation performance. To obtain a more reliable estimate of model performance, reduce bias from a single train-test split, and address the mild class imbalance in the data, we used both stratified and standard k-fold cross-validation with k=5, which is suitable for a moderate size dataset. During model training, the training/validation dataset created as described above was divided into five equal partitions. Then, in each iteration, one partition was used for validation, while the remaining four partitions were used to train the models. This allowed 80% of the training data to be used for model training and the remaining 20% for validation [51]. This process was repeated five times so that each partition was used once for validation. A graphical illustration of the 5-fold cross-validation procedure is shown in Fig. 4.

**Fig 4.**
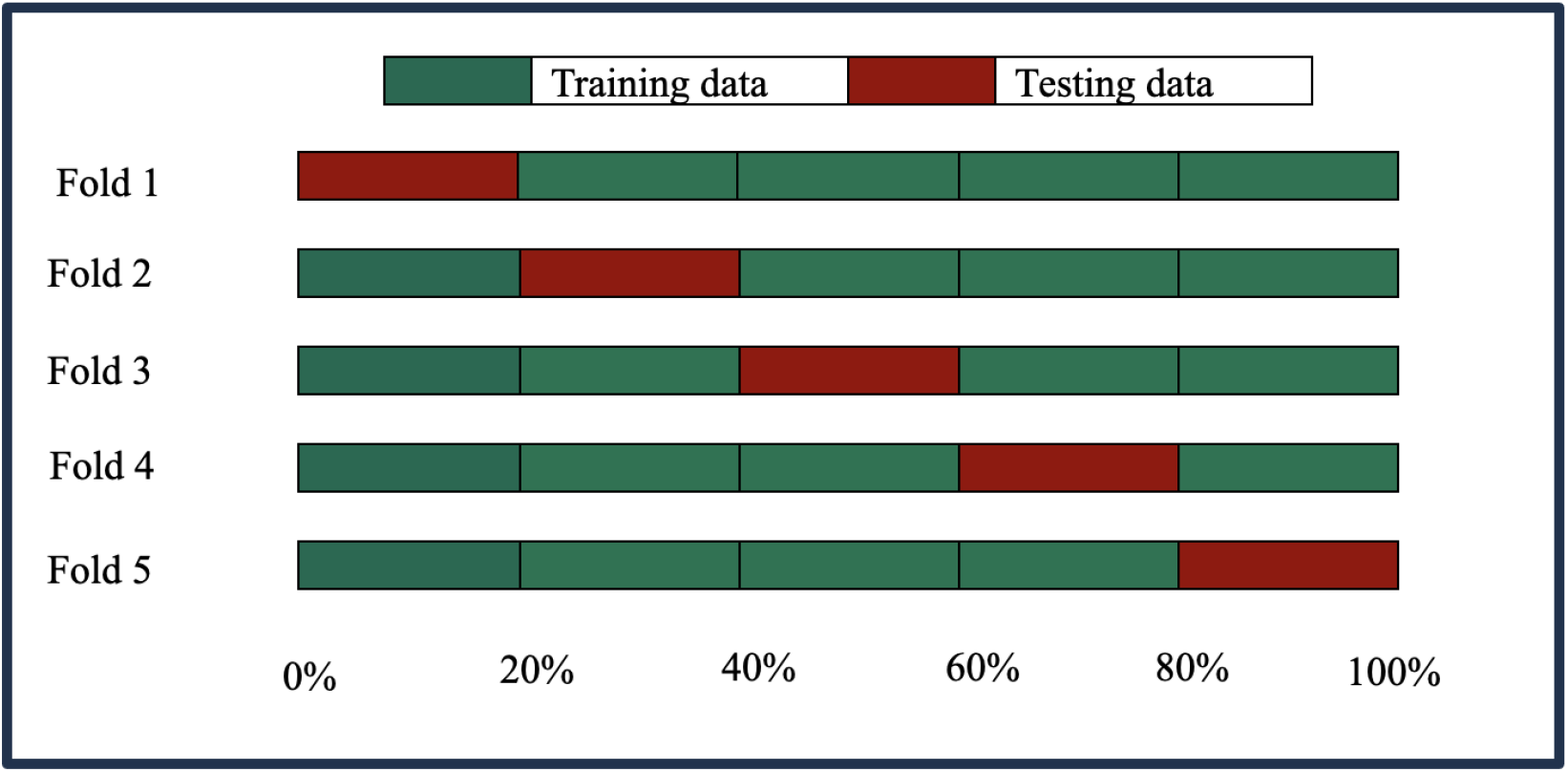
Segregation frame of training and test data used in k-fold cross-validation scheme.

The stratified 5-fold cross-validation procedure is similar to the random 5-fold cross-validation described above, except that, during the partitioning of the training data, care is taken to ensure that the class distribution in each fold matches that of the original training/validation dataset. A graphical illustration of the stratified 5-fold cross-validation procedure is shown in Fig. 5.

**Fig 5.**
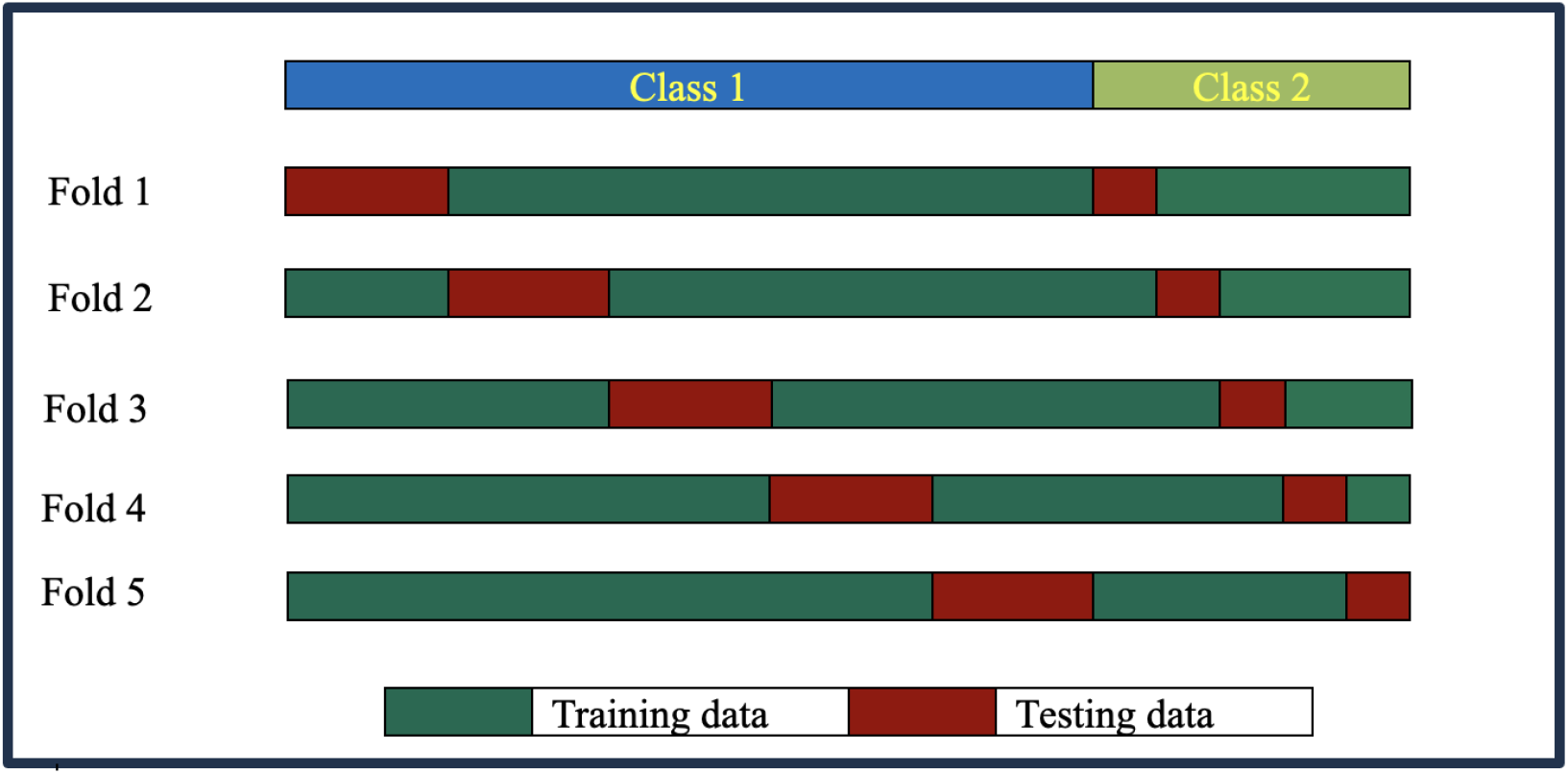
Segregation frame of training and test data used in stratified k-fold cross-validation scheme.

The model development procedure began with data preprocessing, and the dataset used in this study shows a wide range of values, as indicated by the descriptive statistics presented in Table 1. Therefore, to prevent the dominance of one or more input features, data preprocessing is essential. Consequently, the feature space was rescaled using MinMax normalization, which helps achieve optimal model performance. This preprocessing step was carefully applied within each cross-validation fold using a pipeline approach to ensure that no data leakage occurred.

All machine learning models described in the supplementary S1 File materials were trained using the GridSearchCV technique to identify optimal hyperparameters for each model, with MinMax scaling embedded within the pipeline to ensure leak-free tuning across all folds. The hyperparameter search spaces for each model are as follows. For SVM, the parameter space includes the penalty parameter C = {0.01, 0.1, 1, 10, 15, 20, 30, 50, 100, 1000} and kernel functions {linear,rbf, sigmoid}. For KNN, the number of nearest neighbors ranges from 1 to 20, with p = {1, 2, 3}. For RF, the parameter space includes the number of trees {5, 10, 15, 20, 25, 50, 100, 150}, maximum features {sqrt, log2, None}, maximum depth {1, 2, 3, 6, 9}, and maximum leaf nodes {3, 6, 9}. For DT, the parameter space includes maximum depth {2, 3, 5, 10, 20}, minimum samples per leaf {5, 10, 20, 50, 100}, and splitting criteria {gini, entropy}. For XGBoost, the parameter space includes learning rate {0.001, 0.01, 0.1}, subsample ratio {0.5, 0.7, 1}, and maximum tree depth {3, 5, 7}. For the logistic regression model, only the regularization parameter was tuned.

Table 3 presents the optimal hyperparameters for each model and their performance under 5-fold stratified cross-validation, while Table 4 presents the corresponding results for standard 5-fold cross-validation.

**Table 3.** Evaluation Metrics for Proposed Models with Stratified 5-Fold Cross Validation with 95% CI.

| Model | Tuned Hyperparameters | AUC | Accuracy | F-1 Score | Precision | Recall |
| --- | --- | --- | --- | --- | --- | --- |
| **RF** | max-d=9, max-ln=9, n-est= 100 | **0.979**<br>[0.956,0.999] | **0.944%**<br>[0.899,0.989] | **0.959**<br>[0.926,0.992] | **0.960**<br>[0.917,0.999] | **0.959**<br>[0.927,0.991] |
| **SVM** | C=20, kernel='linear' | **0.989**<br>[0.977,0.999] | **0.968%**<br>[0.938,0.997] | **0.976**<br>[0.954,0.998] | **0.976**<br>[0.957,0.995] | **0.976**<br>[0.944,0.999] |
| **DT** | criterion=gini, max-d=3, min-sl=10 | **0.970**<br>[0.948,0.993] | **0.938%**<br>[0.897,0.978] | **0.954**<br>[0.925,0.983] | **0.950**<br>[0.904,0.996] | **0.959**<br>[0.921,0.998] |
| **KNN** | n-neighbors=8, p=3 | **0.968**<br>[0.942,0.994] | **0.958%**<br>[0.919,0.997] | **0.969**<br>[0.941,0.998] | **0.970**<br>[0.933,0.999] | **0.969**<br>[0.942,0.997] |
| **LR** | C= 10 | **0.987**<br>[0.971,0.999] | **0.954%**<br>[0.901,0.999] | **0.966**<br>[0.928,0.999] | **0.957**<br>[0.907,0.999] | **0.976**<br>[0.944,0.999] |
| **XGBoost** | learning-r=0.01, max-d=3, ss=0.7 | **0.978**<br>[0.951,0.999] | **0.942%**<br>[0.898,0.987] | **0.958**<br>[0.926,0.989] | **0.954**<br>[0.907,0.999] | **0.963**<br>[0.935,0.990] |

Table notes: max-d=max-depth, max-ln=max-leaf-nodes, n-est=n-estimators, min-sl=min-samples-leaf. learning-r=learning-rate,ss=subsample.

**Table 4.** Evaluation Metrics for Proposed Models with Random 5-Fold Cross Validation with 95% CI.

| Model | Tuned Hyperparameters | AUC | Accuracy | F-1 Score | Precision | Recall |
| --- | --- | --- | --- | --- | --- | --- |
| **RF** | max-d=9, max-ln=9, n-est=50 | **0.981**<br>[0.961,0.999] | **0.944%**<br>[0.918,0.970] | **0.958**<br>[0.935,0.981] | **0.952**<br>[0.914,0.990] | **0.966**<br>[0.933,0.998] |
| **SVM** | C=5, kernel='linear' | **0.988**<br>[0.979,0.997] | **0.963%**<br>[0.947,0.979] | **0.971**<br>[0.956,0.988] | **0.972**<br>[0.945,0.999] | **0.972**<br>[0.948,0.996] |
| **DT** | criterion=gini, max-d=3, min-sl=10 | **0.969**<br>[0.942,0.998] | **0.944%**<br>[0.904,0.985] | **0.958**<br>[0.924,0.992] | **0.949**<br>[0.887,0.999] | **0.969**<br>[0.950,0.988] |
| **KNN** | n-neighbors=7, p=3 | **0.969**<br>[0.947,0.992] | **0.958%**<br>[0.937,0.980] | **0.969**<br>[0.949,0.987] | **0.968**<br>[0.937,0.999] | **0.969**<br>[0.936,0.999] |
| **LR** | C=10 | **0.986**<br>[0.975,0.998] | **0.956%**<br>[0.928,0.984] | **0.967**<br>[0.942,0.992] | **0.962**<br>[0.917,0.999] | **0.972**<br>[0.948,0.996] |
| **XGBoost** | learning-r=0.1, max-d=3, ss=0.5 | **0.979**<br>[0.959,0.998] | **0.951%**<br>[0.909,0.993] | **0.963**<br>[0.928,0.997] | **0.967**<br>[0.917,0.999] | **0.958**<br>[0.926,0.991] |

Table notes: max-d=max-depth, max-ln=max-leaf-nodes, n-est=n-estimators, min-sl=min-samples-leaf. learning-r=learning-rate,ss=subsample.

Although the data set exhibits a mild class imbalance between PD patients and healthy individuals, both methods, stratified 5-fold cross-validation and random 5-fold cross-validation, produced similar results across all performance metrics. These results suggest that the classification models are relatively stable and are not highly affected by mild class imbalance within the folds. In our study, models trained under stratified 5-fold cross-validation are more complex compared to models developed using standard random 5-fold cross-validation. Furthermore, both methods produce very similar evaluation metrics, such as Precision, Recall, and other evaluation metrics, while maintaining lower complexity. Stratification produces more complex models without providing any greater improvements in predictive power. Given the absence of significant performance gains and the preference for more parsimonious models with better generalization potential, the random 5-fold cross-validation approach was selected for model training and evaluation. Finally, we used 20% of our test data to validate the performance of the proposed models for unseen data. The results given in Table 5 confirm the high quality of the performance of the proposed models for unseen data.

**Table 5.** Model Performance on Unseen Test Data.

| Model | AUC | Accuracy | F-1 Score | Precision | Recall |
| --- | --- | --- | --- | --- | --- |
| Support Vector Machine(SVM) | **0.965** | **96%** | **0.96** | **0.95** | **0.96** |
| Random Forest(RF) | 0.942 | 94% | 0.93 | 0.94 | 0.93 |
| Logistic Regression(LR) | 0.924 | 94% | 0.93 | 0.94 | 0.92 |
| K-Nearest Neighbor(KNN) | 0.927 | 94% | 0.92 | 0.92 | 0.93 |
| Decision Tree(DT) | 0.942 | 94% | 0.93 | 0.93 | 0.94 |
| Gradient Boosting(XGBoost) | 0.958 | 95% | 0.95 | 0.94 | 0.96 |

## 4 Discussion and Conclusion

In this study, our main objective is to enhance the classification of patients with PD from healthy individuals utilizing six well-known AI-based models: Support Vector Machine(SVM), Decision Tree(DT), Logistic Regression(LR), K-Nearest Neighbor(KNN), XGBoost, and Random Forest(RF), each discussed in the supplementary materials. Each of these models excels at identifying nonlinear relationships and interactions among individual risk factors by highlighting critical predictors associated with disease classification. Furthermore, we compared the performance and efficiency of the proposed models using well-defined evaluation metrics such as AUC, Accuracy, F-1 score, Precision, and Recall.

Fig 6 is a graphical visualization of the performance of the six proposed machine learning models under stratified 5-fold cross-validation, and Fig 7 is a graphical illustration of their performance under random 5-fold cross-validation. Fig 6 and 7 show that the SVM model outperforms the remaining machine learning models in all five evaluation metrics for stratified and random 5-fold cross-validation.

**Fig 6.**
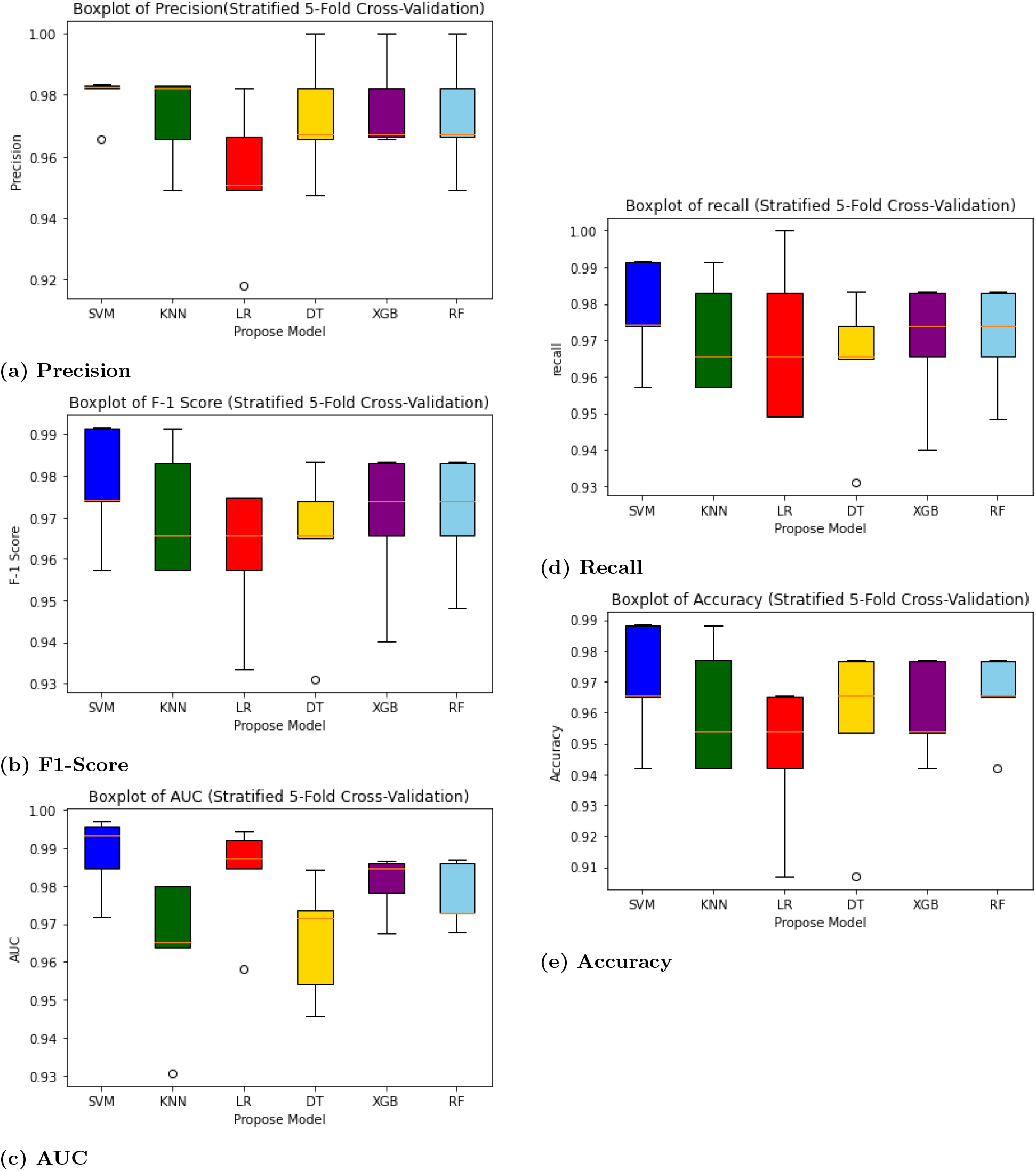
Evaluation of Proposed Models Over Evaluation Metrics Performance with Stratified 5-fold Cross Validation

**Fig 7.**
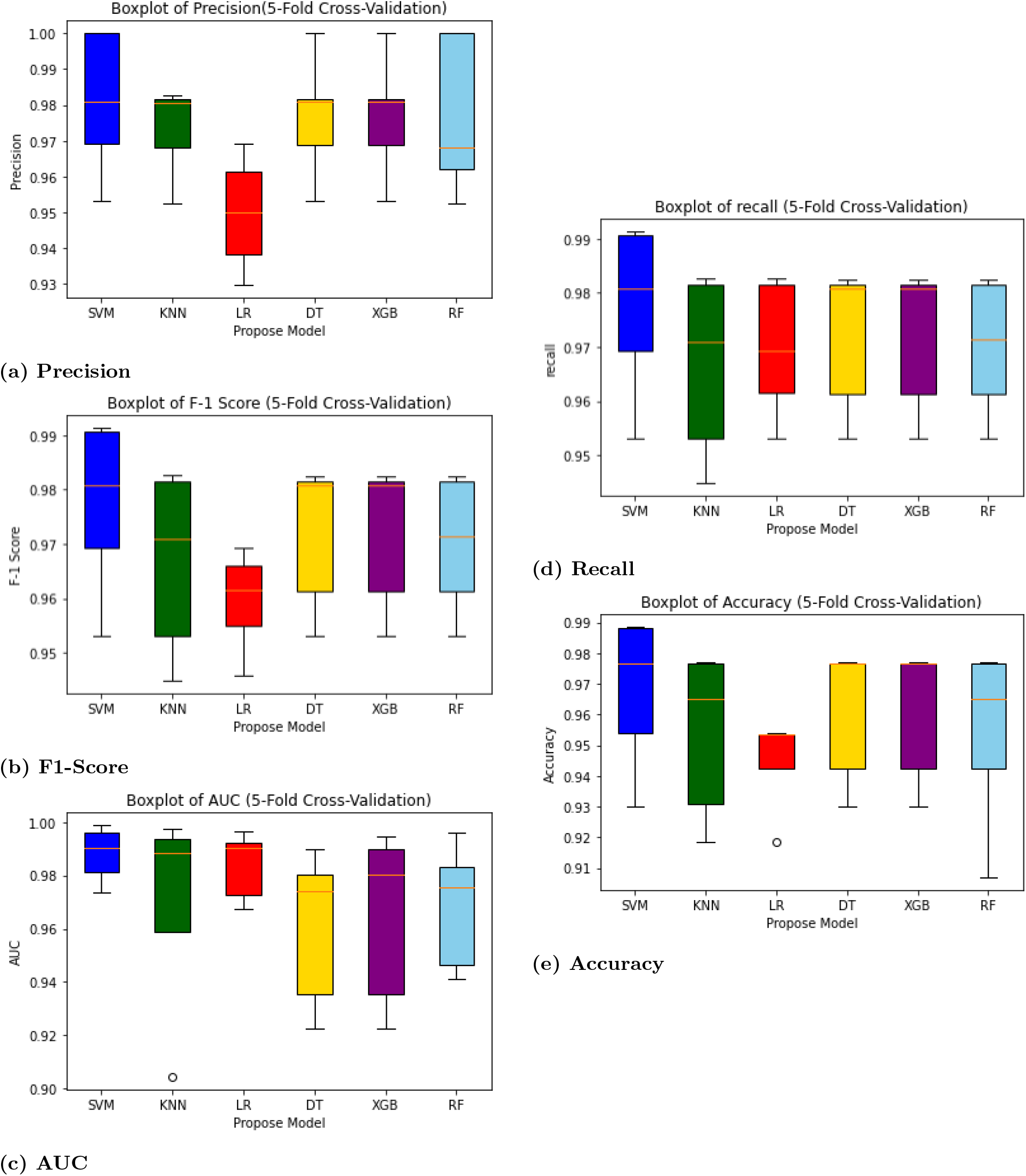
Evaluation of Proposed Models Over Evaluation Metrics Performance with 5-fold Cross Validation

Furthermore, the proposed SVM model also performed better in the test data, as shown in Table 5 in Section 3, confirming its ability to generalize to unseen data. Additionally, our study employed SHapley Additive exPlanations (SHAP) to enhance the interpretability of the final model by identifying the importance of individual risk factors. The SHAP plot in Fig 8 shows the impact of key features in the discrimination of healthy individuals from Parkinson’s patients. PUTAMEN-L and PUTAMEN-R are the leading predictors, where lower values are associated with a higher discriminating power for patients with PD, which aligned with clinical expectations. In contrast, CAUDATE-L with higher values is associated with a higher discrimination power for patients with PD. Other influential features, such as ptau and abeta, indicate higher discrimination power for PD patients when their concentrations are lower. In contrast, higher concentrations of alpha-synuclein(asyn) show stronger predictions of PD. This plot highlights how each factor shapes the classification, emphasizing the importance of brain protein involvement and the behavior of putamen and caudates in classification modeling.

**Fig 8.**
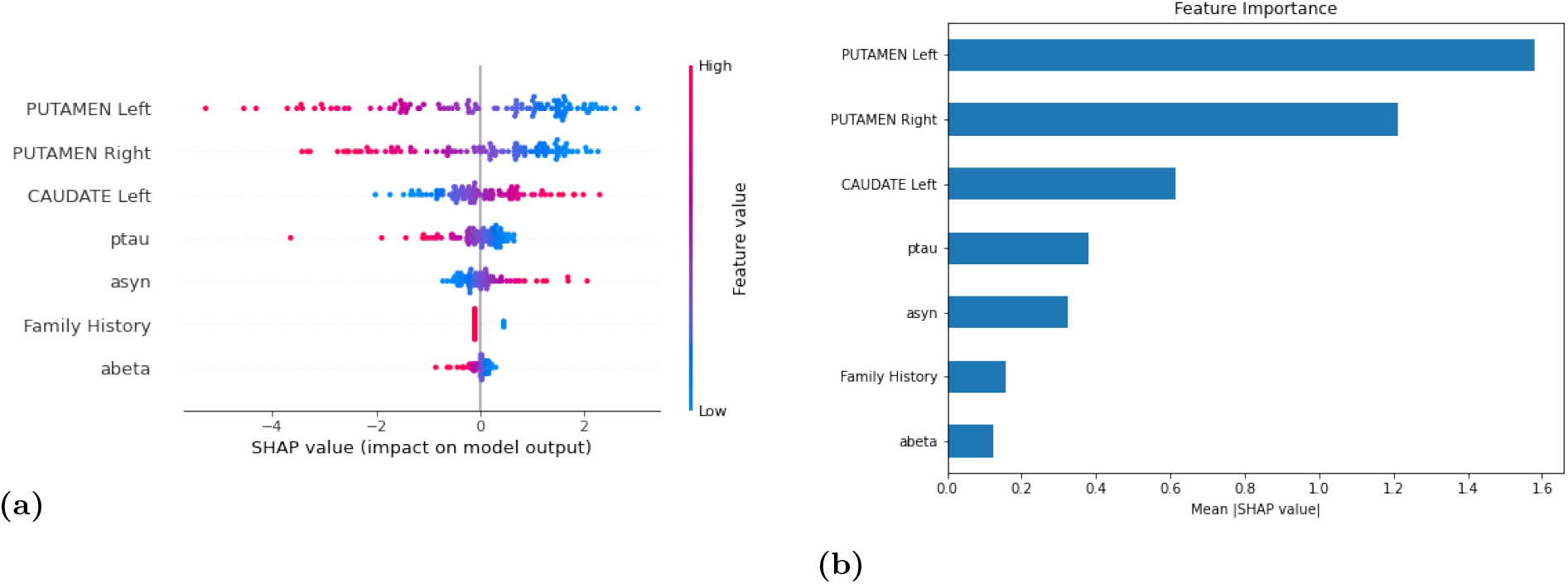
Feature Importance and Beeswarm Plot Determined by Mean Absolute SHAP Values for the SVM model with a Linear Kernel. (a) A comprehensive view of each influence risk factor has on model predictions, with the left putamen ranked on top. (b). Bar plot of mean absolute SHAP values by decreasing order of importance.

PD is a widespread neurological condition that progressively affects the lives of millions of people worldwide, making it one of the most significant public health challenges of our time. It is crucial to have a well-defined formal approach to diagnose patients with PD efficiently and with a high degree of accuracy, rather than relying on descriptive measurements, which helps medical professionals understand the disease and treat it with greater confidence. In this study, we addressed the following:

1. We developed classification models using six widely used machine learning algorithms to identify patients with PD with a high degree of accuracy(at least 98%).
2. The six proposed models were evaluated for their accuracy and efficiency, and their results were validated using well-known robust statistical measurements.
3. The six proposed models were compared using well-defined robust statistical metrics, and the SVM model was identified as the best-performing model to differentiate PD patients from healthy individuals.
4. Finally, this study used SHAP to increase the interpretability of the proposed model and identified the risk factors that contribute the most to the classification of patients with PD.

The robust proposed model utilized seven individual risk factors, and we have identified the importance of these variables in identifying patients with PD. So, professionals may give higher weight to these variables when identifying PD. This might support as a decision-making tool in the early identification of Parkinson’s disease. However, further validation on larger and more diverse real datasets is essential before clinical application. Furthermore, this study contributes to the integration of artificial intelligence in healthcare, emphasizing the value of data-driven classification modeling techniques in supporting healthcare professionals with accurate, personalized, and actionable insights for high-risk patients. Together, these approaches enhance the precision of early PD detection, paving the way for more informed clinical decision-making and improved patient care.

In the literature, we found several articles that utilized individual/ several machine learning techniques to detect PD. So in this paper, we trained, evaluated, and compared widely used machine learning classification models using an interpretable heterogeneous feature set to identify a robust machine learning model that discriminates PD patients from healthy individuals with a high degree of confidence. Despite these promising results in detecting patients with Parkinson’s Disease (PD) at an early stage, this study has some limitations. First, the available PD datasets are relatively small, and early-stage or atypical cases are underrepresented. In addition, the data used in this study were obtained from a single source, which may have reduced the model’s robustness when applied to new or unseen data. Furthermore, biomarkers such as putamen and caudate measurements, *α*-synuclein (*α*-syn), phosphorylated tau(ptau), and amyloid-beta levels can vary significantly across institutions. These differences often arise from variations in scanner settings, sensor configurations, and recording environments. As a result, the model may struggle to generalize well and consistently identify disease patterns across different datasets. Another limitation is the exclusion of genetic, molecular, and lifestyle factors, which may provide important insights into disease progression.

Future studies should focus on collecting data from multiple locations, data centers, and countries, incorporating diverse populations and data types. Exploring multi-modal approaches that integrate imaging, genomic information, and relevant clinical variables could significantly improve predictive accuracy, generalizability, and clinical usefulness. With richer and more diverse data integration, the proposed approach has strong potential to translate research findings into clinical applications, ultimately improve the risk assessment, enabling more personalized treatment strategies, and enhance patient outcomes for individuals with PD.

## Data Availability

Data used in the preparation of this article was obtained on [2021-10-18] from the Parkinson's Progression Markers Initiative (PPMI) database (www.ppmi-info.org/accessdataspecimens/ download-data), RRID: SCR˙006431. For up-to-date information on the study, visit www.ppmi-info.org.

## Supporting information

**S1 File. Machine Learning Classification Models**.

## Acknowledgments

PPMI – a public-private partnership – is funded by the Michael J. Fox Foundation for Parkinson’s Research and funding partners, including 4D Pharma, Abbvie, AcureX, Allergan, Amathus Therapeutics, Aligning Science Across Parkinson’s, AskBio, Avid Radiopharmaceuticals, BIAL, BioArctic, Biogen, Biohaven, BioLegend, BlueRock Therapeutics, Bristol-Myers Squibb, Calico Labs, Capsida Biotherapeutics, Celgene, Cerevel Therapeutics, Coave Therapeutics, DaCapo Brainscience, Denali, Edmond J. Safra Foundation, Eli Lilly, Gain Therapeutics, GE HealthCare, Genentech, GSK, Golub Capital, Handl Therapeutics, Insitro, Jazz Pharmaceuticals, Johnson & Johnson Innovative Medicine, Lundbeck, Merck, Meso Scale Discovery, Mission Therapeutics, Neurocrine Biosciences, Neuron23, Neuropore, Pfizer, Piramal, Prevail Therapeutics, Roche, Sanofi, Servier, Sun Pharma Advanced Research Company, Takeda, Teva, UCB, Vanqua Bio, Verily, Voyager Therapeutics, the Weston Family Foundation and Yumanity Therapeutics.

## Notes

### Competing Interest Statement

The authors have declared no competing interest.

### Summary of Updates

The entire manuscript was updated to improve clarity, correct typographical errors, update references based on peer-reviewed articles, and enhance the overall flow and coherence.

