## Supplementary Document S1 File for "Data-driven robust machine learning models to differentiate Parkinson’s disease patients using heterogeneous risk factors"

### 1 Machine Learning Classification Models

In recent years, the amount of data has increased rapidly, creating a strong need for simple and efficient ways to analyze it. Artificial intelligence (AI), especially machine learning, is widely used to study data in many fields, including healthcare. However, in healthcare, collecting large datasets can be difficult because of privacy concerns and high costs. Even with these challenges, supervised machine learning methods can still work well with smaller datasets. In this study, our main goal is to classify individuals as Parkinson's disease (PD) or as healthy individuals. To do this, we use six common supervised machine learning methods: Random Forest, Extreme Gradient Boosting (XGBoost), Support Vector Machine, Decision Tree, K-Nearest Neighbors, and Logistic Regression. We compare these methods to find the most accurate and reliable approach. This supplementary material provides a brief overview of each of these six models used in our study.

#### 1.1 Decision Tree (DT)

A Decision Tree (DT) is a supervised machine learning algorithm used for both classification and regression tasks. It starts with a root node that represents the entire dataset and splits the data into smaller, more uniform groups based on selected features and thresholds. This process continues step by step, creating child nodes, until a stopping condition is reached, such as when all data in a node belongs to the same class or a maximum depth is reached. The final nodes, called leaf nodes, provide the predicted outcomes or class labels. The splits are determined using measures such as information gain, Gini impurity, or variance reduction. The performance of a decision tree depends on factors like data quality, tree depth, splitting method, and pruning, with clean and well-prepared data generally leading to better accuracy. More details on DT, splitting rules, and tree pruning methods can be found in the literature, [1], [2]

### 1.2 Random Forest (RF)

Random Forest (RF) is a well-known supervised machine learning technique that has been successfully used for feature selection and statistical modeling tasks such as classification and regression. RF, first introduced by L. Breiman, multiple decision trees can be integrated into a single model using the concept of ensemble learning, and this combination of trees is defined as a Random Forest, [3], [4], [5]. The Random Forest technique trains several decision trees using bootstrap resampling and combines their decisions to produce the final outcome. In the Random Forest model, the bootstrap resampling method is applied to create sub-samples of the training dataset with replacement. Separate decision trees are then built for each sub-sample, referred to as weak learners, and these weak learners are combined to build one strong learner, known as the Random Forest model. For regression tasks, the predictions of each decision tree are averaged to obtain the final output. For classification tasks, the majority decision among the individual decision trees is taken as the final decision of the Random Forest model, [6], [7]. There are several advantages of using RF over DT. For example, RF uses ensemble learning, which reduces the risk of overfitting. Additionally, bootstrapping allows RF to work effectively with small datasets, and RF has been successfully applied in medical research, [8], [9]. Figure 1 illustrates the basic underlying idea of RF and the final decision-making process for classification tasks.

### 1.3 Extreme Gradient Boost (XGBoost)

Gradient Boosting is a special type of machine learning ensemble model that is used for Classification and Regression tasks (CART tree) and was introduced by Friedman, [10]. XGBoost was first introduced by Tianqi Chen and Carlos Guestrin in 2016, and the main idea behind XGBoost is to iteratively combine and train a number of weak learners to develop one strong learner, [11], [12]. This weak learner combination method is known as ensemble learning, a powerful supervised machine learning technique. In the XGBoosting approach, some models correctly predict the task, but some are weak learners, meaning they do not

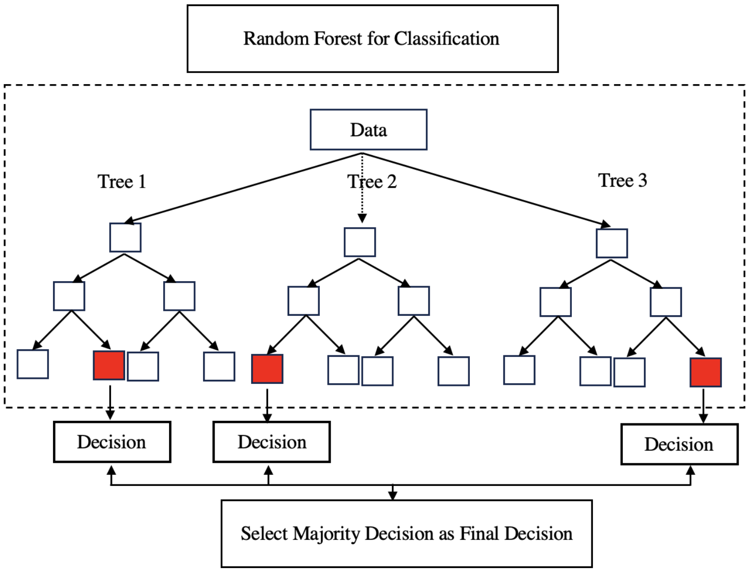


Figure 1: Random Forest Classification

predict the required task correctly. Thus, in the next iteration, more weights will be assigned to the weak learning models that incorrectly classify the output task than the models that correctly classify the output, [11]. The following section briefly explains the training process of the XGBoost model.

Suppose we have a Classification and Regression (CART) task with $n$ number of training examples, say, $(x_1, y_1)$, $(x_2, y_2)$..., $(x_n, y_n)$. Then $n$ number of CART trees were created and combined by utilizing the ensemble learning technique to integrate a single strong, efficient model. Thus, the predicted output of each CART tree is $\hat{y}_i = f_i(x_i)$, and the output of the ensemble final model can be defined as follows,

$$\hat{y} = \sum_{i=1}^{n} f_i(X_i), \qquad f_i \in \zeta \tag{1}$$

where $\zeta$ is the space of all CART.

The main objective of the training process is to minimize the objective function of the proposed model, which is defined by,

$$obj(\theta) = \sum_{i=1}^{n} l(\hat{y}_i, y_i) + \sum_{i=1}^{n} \beta(f_i), \tag{2}$$

where $l(\hat{y}_i, y_i)$ is the training loss for the individual CART trees, and this can be defined as the difference between the predicted output and the actual output of the individual CART trees. However, when it comes to the regression trees, the Mean Squared Error (MSE) loss function is used as the loss function, and it is given by the following Equation 3,

$$MSE = \sum_{i=1}^{n}(y_i - \hat{y_i})^2. \tag{3}$$

On the other hand, for the classification models, the logistic loss is the loss function on the training data, given by Equation 4,

$$\text{Logistic Loss} = \sum_{i=1}^{n}[y_i ln(1 + e^{-\hat{y_i}}) + (1 - y_i) ln(1 + e^{-\hat{y_i}})]. \tag{4}$$

The second part of Equation 2 is called the regularization part of the XGBoost model, and can be defined as Equation 5. This regularization part helps the XGBoost model to penalize the complexity. Thus, the XGBoost model avoids the overfitting effect through regularization.

$$\beta(f_i) = \gamma T + \frac{1}{2}\lambda \|W^2\|, \tag{5}$$

where $T$ is the total number of leaves, $W$ is the leaf score, $\lambda$ is the regularization parameter, and $\gamma$ is the information gain.

XGBoost model consists of several hyperparameters such as the learning rate, the maximum depth that the tree allows to grow (max-depth), the number of trees in the model (n-estimators), the sub-sample size, the information gain (gamma), and the regularization parameter. These hyperparameters need to be predefined before starting the training procedure, and during the training process, the developer can tune these hyperparameter values to identify their optimal values for the final model, [13], [14]. In this study, we utilize GridSearchCV to tune predefined hyperparameters.

Once we finished the hyperparameter tuning process and finalized the developed XGBoost model, it provided a score for each feature, indicating the importance of each feature in the proposed model. Those scores are used to understand how important each variable is when we are performing the proposed classification task. There are three types of feature importance, and they can be defined in the following ways,

- **Gain:** A metric of how much the objective function could be improved by adding a branch of the regression tree with a given function.
- **Weight:** It represents the number of features that are used by all the regression trees.
- **Cover:** It is the average of the number of times a single input is branched by a feature.

### 1.4 Support Vector Machine (SVM)

SVM is one of the powerful state-of-the-art machine learning techniques that has been utilized to solve problems in various domains such as Health Sciences, Finance, Environmental Sciences, etc. In 1995, Vladimir Vapnik introduced SVM to analyze classification and regression tasks, [15]. To illustrate mathematical intuition beneath SVM, suppose we have a training data set that consists of explanatory variables and the target variable, which is a binary variable with a positive class and a negative class (i.e, $y \in \{-1, 1\}$). Let's consider a hyperplane ($H_1$) that separates the positive class from the negative class for a given set of training data, as illustrated in Figure 2.

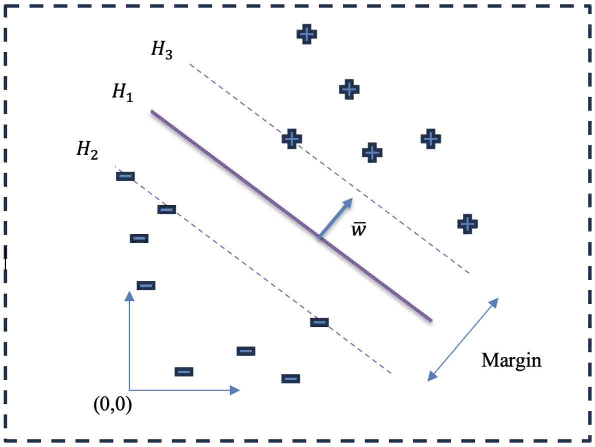


Figure 2: Linearly Separable Hyperplane 1

Assume that the points on the hyperplane ($H_1$) satisfy $\bar{w}.\bar{x} + b = 0$, where $\bar{w}$ is perpendicular to the hyperplane, as shown in Figure 2. Suppose that all training examples satisfy the following constraints,

$$\bar{w}.\bar{x} + b \geq 1, \quad if \quad y_i = +1\,(Positive\,class) \tag{6}$$

$$\bar{w}.\bar{x} + b \leq -1, \quad if \quad y_i = -1\,(Negative\,class) \tag{7}$$

Let's define the target variable in the training data as follows,

$$y_i = \begin{cases} 1, & for \quad Positive\,class \\ -1, & for \quad Negative\,class \end{cases}$$

Thus, inequalities given by Equation 6 and Equation 7 can be summarized as follows,

$$y_i(\bar{w}.\bar{x} + b) - 1 \geq 0. \tag{8}$$

The points $x_1$ and $x_2$ on the hyperplanes $H_3$ and $H_2$ must satisfy the following relationships, respectively.

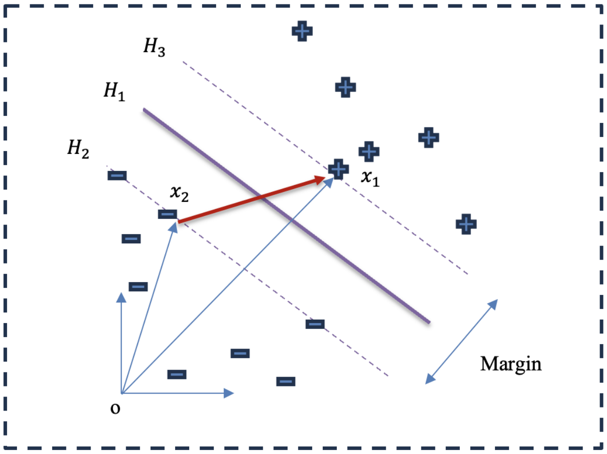


Figure 3: Linearly Separable Hyperplane 2

$$(\bar{w}.\bar{x_2} + b) - 1 = 0, \quad y_i = 1 \tag{9}$$

$$(\bar{w}.\bar{x_1} + b) + 1 = 0, \quad y_i = -1 \tag{10}$$

For given vectors $\overrightarrow{ox_1}$, $\overrightarrow{ox_2}$ and $\overrightarrow{x_2x_1}$, following is true,

$$\begin{aligned} \overrightarrow{ox_2} &= \overrightarrow{ox_1} + \overrightarrow{x_2x_1} \\ \overrightarrow{x_2x_1} &= \overrightarrow{ox_1} - \overrightarrow{ox_2} \end{aligned} \tag{11}$$

From Equations 9, 10 and 11, we have $\overrightarrow{x_2x_1}$,

$$\begin{aligned} \overrightarrow{x_2x_1} &= (\bar{w}.\bar{x_1} + b) + 1 - (\bar{w}.\bar{x_2} + b) + 1 \\ &= \bar{w}.(\bar{x_2} - \bar{x_1}) + 2. \end{aligned} \tag{12}$$

Now, the width between $H_2$ and $H_3$, which is the margin, can be derived using the vector dot product,

$$H_2H_3 = \overrightarrow{x_2x_1}.\frac{\overrightarrow{w}}{||w||}. \tag{13}$$

From Equations 12 and 13 the following is true,

$$H_2H_3 = \frac{2}{||w||}. \tag{14}$$

The main objective of SVM is to maximize the margin or distance between $H_2$ and $H_3$ subject to the constraints given by Equation 8. Thus, according to Equation 12, $||w||$ should be minimized. For mathematical simplicity let's consider $\frac{1}{2} * ||w||^2$ and minimize instead of $||w||$ subject to the constraint defined by the equation 8. Thus, we utilize Lagrangian multipliers as given below,

$$L = \frac{1}{2} * ||w||^2 - \sum \lambda_i [y_i(\bar{w}.\bar{x} + b) - 1]. \tag{15}$$

Using Equation 15, the following is true,

$$\frac{\partial L}{\partial w} = \bar{w} - \sum \lambda_i y_i.\bar{x},$$

$$\bar{w} = \sum \lambda_i y_i.\bar{x}. \tag{16}$$

$$\frac{\partial L}{\partial b} = -\sum \lambda_i.y_i$$

$$0 = \sum \lambda_i.y_i \tag{17}$$

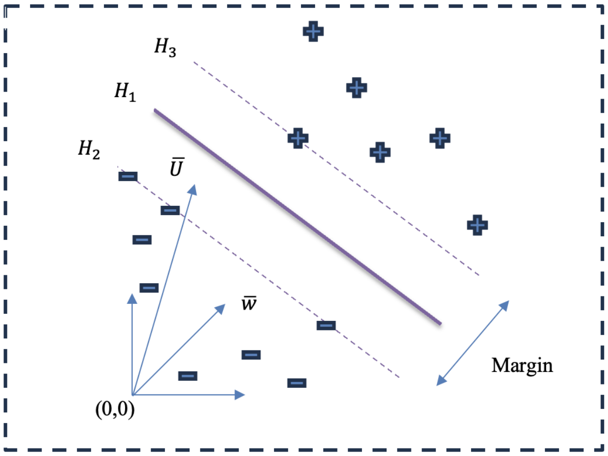


Figure 4: Linearly Separable Hyperplane 3

### 1.5 K-Nearest Neighbor (KNN)

KNN (K-Nearest Neighbors) is a simple non-parametric supervised machine learning algorithm used for both classification and regression. It was first introduced by Evelyn Fix and

Joseph Hodges in 1951 and later improved by Thomas Cover in 1967. The main idea behind KNN is that a new data point can be classified based on the characteristics of nearby data points. To do this, KNN measures the distance between the new data point and existing points, then selects the closest neighbors. The new point is assigned to the class that appears most often among these neighbors. Common distance measures used in KNN include Euclidean, Manhattan, Minkowski, and Hamming distances. Another important factor in KNN is the number of neighbors, called K. This value determines how many nearby points are considered when making a decision. In most cases, K is chosen as an odd number to reduce the chance of ties between classes. More detailed explanations of KNN can be found in prior research studies, [16], [17], [18]..

### 1.6 Logistic Regression (LR)

LR is a well-known supervised machine learning algorithm used to accomplish dichotomous classification tasks by predicting the probability of model outcomes in various domains such as medical diagnosis, sentiment analysis, and environmental sciences, among others. The LR model is used to model the log odds of an event as a linear combination of one or more input risk factors. Let us consider $x_1$,$x_2$,...$x_n$ as input risk factors. Then the analytical form of the Logistic Regression model is given by,

$$log\left(\frac{p}{1-p}\right) = \alpha_0 + \alpha_1 x_1 + \alpha_2 x_2 + ... + \alpha_n x_n, \tag{18}$$

where p is the probability of the occurrence of the event of interest, $x_i$s are individual risk factors, $\alpha_0$ is the bias term, and $\alpha_i$s are the weights of the individual risk factors. Finally, we utilize the logistic function, commonly known as the sigmoid function, to convert the model output, given by Equation 18, into its representative probability value. The general form of the sigmoid function is given by,

$$p(x) = \frac{1}{1+e^{-z}}, \tag{19}$$

where $p(x)$ is the probability of the event of interest, $z = \alpha_0 + \alpha_1 x_1 + \alpha_2 x_2 + .... + \alpha_n x_n$, $x_i$s are individual risk factors, $\alpha_i$s represent the weights of the individual risk factors, and $\alpha_0$ is the bias term. If the probability is less than a predefined threshold (0.5), we predict the response as 0, and if the probability is greater than or equal to that predefined threshold (0.5), we predict the response as 1. Further details on Logistic Regression can be found in the literature, [17].

[3] Leo Breiman. Random forests. *Machine Learning*, 2001.

[4] Rohini Srivastava, Shailesh Kumar, and Basant Kumar. 7 - classification model of machine learning for medical data analysis. In Tilottama Goswami and G.R. Sinha, editors, *Statistical Modeling in Machine Learning*, pages 111–132. Academic Press, 2023.

[5] Siddharth Misra and Hao Li. Chapter 9 - noninvasive fracture characterization based on the classification of sonic wave travel times. In Siddharth Misra, Hao Li, and Jiabo He, editors, *Machine Learning for Subsurface Characterization*, pages 243–287. Gulf Professional Publishing, 2020.

[6] Gilles Louppe. Understanding random forests: From theory to practice, 2015.

[7] A. Chaitanya Kumar, J. Andrew John, Maheswari Raja, and P. Vijaya. Chapter 4 - genetic factor analysis for an early diagnosis of autism through machine learning. In Amit Kumar Tyagi and Ajith Abraham, editors, *Data Science for Genomics*, pages 69–84. Academic Press, 2023.

[8] Khalilia M Chakraborty S Popescu M. Predicting disease risks from highly imbalanced data using random forest. *BMC Med Inform Decis Mak*, 2011.

[9] Sarica A Cerasa A Quattrone A. Random forest algorithm for the classification of neuroimaging data in alzheimer's disease: A systematic review. *Front Aging Neurosci*, 2017.

[15] Shmilovici Armin. *Support Vector Machines*. Springer US, Springer US.

[16] Uddin Shahadat Haque Ibtisham Lu Haohui Moni Mohammad Ali Gide Ergun. Comparative performance analysis of k-nearest neighbour (knn) algorithm and its different variants for disease prediction. *Scientific Reports*, 2022.

[17] Lopez-Bernal Diego Balderas David Ponce Pedro Molina Arturo. Education 4.0: Teaching the basics of knn, lda and simple perceptron algorithms for binary classification problems. *Future Internet*, 13, 2021.

[18] Li-Yu Hu Min-Wei Huang Shih-Wen Ke Chih-Fong Tsai. The distance function effect on k-nearest neighbor classification for medical datasets. *SpringerPlus*, 5, 2016.
